# Epigenetic and Immunometabolic Signatures of Suicidal Behavior in Major Depressive Disorder

**DOI:** 10.64898/2026.08.27.26361547

**Authors:** Qiong Sha, Martha L Escobar Galvis, Zach, Zhen Fu, Ryan D. Sheldon, Tyce Cave, Marie Adams, Christine Isaguirre, LeAnn Smart, Janelle Kassien, Timothy Triche, Yvonne Fondufe-Mittendorf, Nagy A. Youssef, Eric D. Achtyes, J. John Mann, Lena Brundin

## Abstract

Suicidal behavior results from complex behavioral and biological changes. Previous crosssectional studies indicate that proinflammatory immunobiological factors are often increased in close temporal proximity to a suicide attempt. Suicidal individuals may also exhibit a biological trait vulnerability to stress and inflammation, due to persistent epigenetic modifications.

We enrolled 130 individuals with major depressive disorder (MDD), 83 with suicidal behavior at intake, and followed them for 12 months with up to eight clinical assessments. Quantification of plasma inflammatory markers and metabolites was performed by high-sensitivity electrochemiluminescence and Ultra High-Performance-Liquid-Mass Spectrometry (UPLC-MS), respectively. Epigenetic changes were identified using Illumina EPIC arrays.

We identified 15 genes with altered DNA-methylation associated with suicidal behavior and attempts at baseline. Childhood trauma predicted lifetime suicide attempts and was associated with altered methylation of seven genes. Increased neutrophils and lower plasma serotonin at baseline predicted future suicide attempts over the following year (neutrophil estimate = 0.42, *P* = 0.016; serotonin OR = 0.58, 95% CI: 0.39–1.13). Utilizing biomarkers from baseline and epigenetic data from the genes with highest predictive values (*STBD1, PRDM8*, and *TRIM15*), we achieved an area under the curve (AUC) of 0.84 for suicide attempts over the year.

Suicidal behavior in MDD was associated with specific epigenetic signatures. Several of the identified genes, such as *MAD1L1*, have been implicated in psychiatric disease, suicidal behavior and the immune response. These findings support the usefulness of epigenetic and immunometabolic blood markers for identifying suicidal individuals in clinical settings, potentially enhancing preventative efforts.

## Introduction

Suicidal behavior constitutes a major cause of mortality and morbidity worldwide, and because it has genetic and epigenetic components, improved understanding of these and other biological factors can enhance its prediction and prevention. We and others have reported that suicide attempts and deaths by suicide are linked to inflammatory activation, detectable in blood, cerebrospinal fluid, and brain tissues (1–3). We have also demonstrated that ongoing inflammation in individuals with suicidal behavior associates with altered production of tryptophan-derived metabolites through the kynurenine pathway (4, 5). Overactivation of this pathway also has the potential to reduce serotonin levels, by diverting tryptophan away from serotonin synthesis (6).

Other biological factors can act as long-term or “trait” regulators of risk for suicidal behavior. Epigenetic modifications, including DNA-methylation, regulate gene transcription and can impact short– and long-term responses to stressors (7, 8). Exposure to traumatic events during childhood is a long-term risk-factor for suicidal behavior. Childhood trauma is linked to increased DNA-methylation of genes involved in stress regulation, including the *NR3C1* gene, encoding the glucocorticoid receptor (9–11). Such epigenetic alterations in childhood or adolescence, especially when related to emotional and physical trauma, can make individuals susceptible to heightened stress responses later in life (12, 13). Some trauma-induced DNA-methylation changes can also persist across generations, affecting both gene expression and the metabolome (14–16), and may therefore contribute to the familial transmission of suicidal behavior alongside genetic inheritance (17).

In this study, we propose that dysregulation of immunometabolic factors associates with suicidal behavior, particularly in individuals with underlying epigenetic vulnerability. To test this hypothesis, we prospectively enrolled subjects with major depressive disorder (MDD), with and without active suicidal behavior, at baseline in a year-long longitudinal clinical study. Blood was collected and participants completed symptom-assessments, trauma– and medical questionnaires at up to eight timepoints over the study period.

## Materials and Methods

### Study design

This study was approved by Van Andel Research Institute (VARI) Institutional Review Board, Grand Rapids, Michigan (protocol N° 19010). Inclusion criteria were: ≥18 years of age, diagnosed with MDD, current, as confirmed by The Structured Clinical Interview for DSM-5 (SCID) interview (18) and English speaking. Exclusion criteria included pregnant or nursing women, subjects with dementia or difficulty understanding study procedures thus lacking capacity to give informed consent, a primary psychiatric diagnosis other than MDD, an active somatic disorder involving the immune system, chronic and systemic immunomodulatory treatment, or individuals undergoing active treatment for cancer.

130 individuals with MDD were enrolled, with the first research visit occurring near the hospital intake for most participants. Participants were followed a year, with research visits approximately every two months. Blood samples from 124 subjects were available from the baseline visit for methylation analysis, of which 123 passed quality-control prior. **Table 1** shows the cohort demographics; medications taken are listed **Supplementary Table 1**.

**Table 1.** Demographics and clinical characteristics of the participants whose blood samples were analyzed in this study (n=123). A “yes” response to C-SSRS item 21, assessed over the 7 days prior to enrollment was recorded as current suicidal behavior at baseline. Percentages are calculated within each demographic category. Some categories from the original demographic form have been merged to avoid too small groups in the data presentation (such as details of marital status/living together as well as individual races represented by a few subjects, are included in the “others” category).

|  |  | Current suicidal behavior at baseline |  |
| --- | --- | --- | --- |
|  |  | Yes (N=82) | No (N=41) |
| **Age; median (IQR)** |  | 28 (14) | 32 (20) |
| **Demographic categories; N(%)** |  |  |  |
| Total |  | 82 (67%) | 41 (33%) |
| Sex | Male | 26 (32%) | 15 (37%) |
|  | Female | 56 (68%) | 26 (63%) |
| Race | White/Caucasian | 69 (84%) | 37 (91%) |
|  | Black/African American | 4 (5%) | 2 (5%) |
|  | Other | 6 (7%) | 1 (2%) |
|  | Not disclosed | 3 (4%) | 1 (2%) |
| Ethnicity | Non-Hispanic | 75 (91%) | 34 (83%) |
|  | Hispanic | 4 (5%) | 7 (17%) |
|  | Not disclosed | 3 (4%) | – |
| Marital status | In a relationship | 27 (33%) | 22 (54%) |
|  | Single | 52 (63%) | 19 (46%) |
|  | Not disclosed | 3 (4%) | – |
| Income | ≤ \$34,000 | 25 (30%) | 16 (39%) |
| | \$34,001to \$70,000 | 27 (33%) | 8 (20%) |
| | \$70,001 to \$120,000 | 17 (21%) | 10 (24%) |
| | ≥ \$120,000 | 9 (11%) | 6 (15%) |
|  | Not disclosed | 4 (5%) | 1 (2%) |
| **Rating scales; median total score (IQR)** |  |  |  |
| HAMD |  | 29 (10) | 24 (10) |
| ACE |  | 4 (5) | 3 (3) |

### Assessment of suicidal behavior

Participants were assessed for suicidality using the Columbia-Suicide Severity Rating Scale (C-SSRS) (19) covering the subjects’ lifetime, the past seven days (denoted as “current”, assessed at each visit), and the period since the last visit (assessed at every visit following the first). We used two measurements of suicidality: ***suicidal behavior***, defined by item 21, as any type of active suicidal behavior, including attempts, aborted attempts and active preparation for an attempt; and a ***suicide attempt***, defined as a self-destructive act with the intent to die, for which items 13 (suicide attempt interrupted by someone other than self) and 16 (suicide attempt, failed) were used. Self-aborted suicide attempts and active preparation were not included.

### Clinical assessments

The SCID module A was administered by a trained specialist at visit one (18) and each enrolled subject met criteria for an ongoing episode of MDD. Upon enrollment, subjects also completed the Hamilton Depression Rating Scale (HAMD) (20) and the Adverse Childhood Experiences (ACEs) assessment (21). Current diagnoses and medications, blood pressure, heart rate, weight and height were recorded together with any ongoing somatic symptoms. Individuals were confirmed to have normal temperature and no symptoms of ongoing infection at each research visit.

### Blood sampling

Blood samples were collected by venipuncture in EDTA tubes, from 7:00 to 11:00 am, and immediately transported to the laboratory on ice. Samples were centrifuged at 700 × g for 15 min at 4 °C, plasma was extracted, aliquoted and immediately frozen at –80 °C until analysis. Whole blood for DNA-methylation analysis was collected in a separate vial and immediately stored at – 80 °C.

### Analysis of immunometabolic factors

Cytokines were measured using the Meso Scale Discovery platform and run on a Sector 600 (Meso Scale Diagnostics, MD, USA), as we previously published (22). Samples were run in duplicate, using mean values for further analysis. Inter-assay coefficients of variation (% CV) were: IL-1β (7.0), IL-6 (14.4), IL-8 (11.6) IL-10 (9.4), and TNF-α (12.6).

Serotonin, tryptophan and kynurenine pathway metabolites were measured at VARI Mass Spectrometry Core (RRID:SCR_024903) as we detailed in (23). Kynurenine pathway-related metabolites (tryptophan: TRP; serotonin: 5HT; kynurenine: KYN; kynurenic acid: KYNA; 3-Hydroxykynurenine: 3HK; quinolinic acid: QUIN; picolinic acid: PIC) were extracted using 90% acetonitrile and quantified using reverse phase ultra high-performance-liquid-chromatography (UHPLC; 1290 Infinity II, Agilent Technologies, CA) coupled to a 6470 Triple quadrupole mass spectrometer (Agilent Technologies). Intra-assay % CV for plasma analytes: TRP 1%, 5-HT 4%, KYN 1%, KYNA 2%, 3-HK 3%, QUIN 2%, PIC 3%.

### DNA methylation profiling and data analysis

Genome-wide DNA-methylation was assessed using Infinium Methylation EPIC Kit V2 (Illumina, CA, USA) at VARI Genomics Core (RRID:SCR_022913), as we detailed in (1). Methylation data were first preprocessed and normalized by Quantile method using minfi (1.52.1) (24). One of the 124 samples was excluded following minfi quality-control based on joint distributions of log-transformed median methylated and unmethylated signal intensities, visualized using plotQC (**Supplementary Fig. 1**). After quality-control and normalization, 900,272 probes (targeting 900,272 CpG sites) remained for downstream analysis. Differentially methylated loci (DML) and regions (DMR) were identified using SeSAMe with a standard quality control pipeline (1.24.0) (25) with a linear model adjusted for sex, age, and BMI within the package, using methylation β-values as outcomes. DMR *P*-values were adjusted for multiple comparisons using the Benjamini-Hochberg false discovery rate (FDR).

### Deconvolution analysis

Deconvolution analysis of the methylation data into six cell-types (CD4 T-cells, CD8 T-cells, natural killer cells, B-cells, monocytes and neutrophils) was performed via FlowSorted.Blood.EPIC (2.10.0) (26).

### Statistical analysis

Methylation data from each CpG site were first transformed into M-values, representing the log_2_-transformed ratio of methylated to unmethylated signal intensities derived from normalized β-values. To compare group differences in genes overlapping with DMRs, the average M-value of CpG sites located within transcription start site (TSS) or enhancer regions was calculated for each gene. The M-values (or their averages from each gene) were used for downstream analysis as they were approximately homoscedastic, and superior for statistical modeling. For data visualization, β-values were used as more biologically interpretable (27).

For deconvolution analysis, the cell-type difference between groups was calculated using Dirichlet regression (0.7.2) (28) with age, sex and BMI as covariates.

Due to potential multicollinearity, the association between suicide and environmental trauma was evaluated by ridge regression logisticRidge function in the R-package “ridge” (3.3) (29) with the four suicide-related questions as outcomes (current and lifetime suicidal behavior; current and lifetime suicide attempt) and trauma-related questions including: ACE1, ACE2, ACE3, ACE4, ACE6, ACE8, ACE9, ACE total score, as covariates, adjusting for sex, age and BMI.

The association analysis between the HAMD item-3 question assessing suicidality, and genes, was conducted via ordinal regression using R-package “ordinal” (2023.12.4.1) (30) adjusting for sex, age and BMI.

For the prediction models, LASSO regression was used to filter common DMLs via “glmnet” (4.1.8) (31). Principal Component Analysis (PCA) was performed on the LASSO filtered CpG sites using R-packages “FactoMineR” (2.11) (32) and “factoextra” (1.0.7) (33).

For ranking the significantly methylated genes, ridge regression was used via R-package “glmnet” (4.1.8) (31). For predicting secondary suicidal outcome, Bayesian logistic regression with a horseshoe prior, well suited for handling sparsity and providing robust predictive estimation [40], was used employing R-package “brms” (2.22.0) (34). M-values of all significantly differentially methylated genes and natural log-transformed biomarkers were used as predictors while future suicide attempts were used as outcome. Areas under the receiver operating characteristic curve (ROC-AUC) and precision-recall curve (PRC-AUC) were calculated using the R-package precrec (version 0.14.4) (35). Leave One Out cross-validation was conducted using R-package “loo” (2.8.0) (36).

## Results

### 1. Altered DNA methylation in patients with suicidal behavior at baseline

To examine differences in DNA-methylation signatures, we conducted differential methylation analyses between subjects with and without suicidal behavior and attempts. We identified differentially methylated CpG loci (DML) and regions (DMRs) from each analysis. 38,965 DMLs and 4 DMRs were derived for current suicidal behavior; 36,055 DMLs and 3 DMRs for lifetime suicidal behavior; 39,579 DMLs and 2 DMRs for current suicide attempt; and 40,189 DMLs and 8 DMRs for lifetime suicide attempt.

Within DMRs, we identified overlapping genes located at TSS or in exons. Detailed DMR information from each analysis is provided in **Figs. 1F** and **2J**. Fifteen genes were associated with suicidal behavior and attempts. Among the 15 genes, *NNAT* and *BLCAP* originated from the same probes and were therefore considered a gene pair.

**Figure 1.**
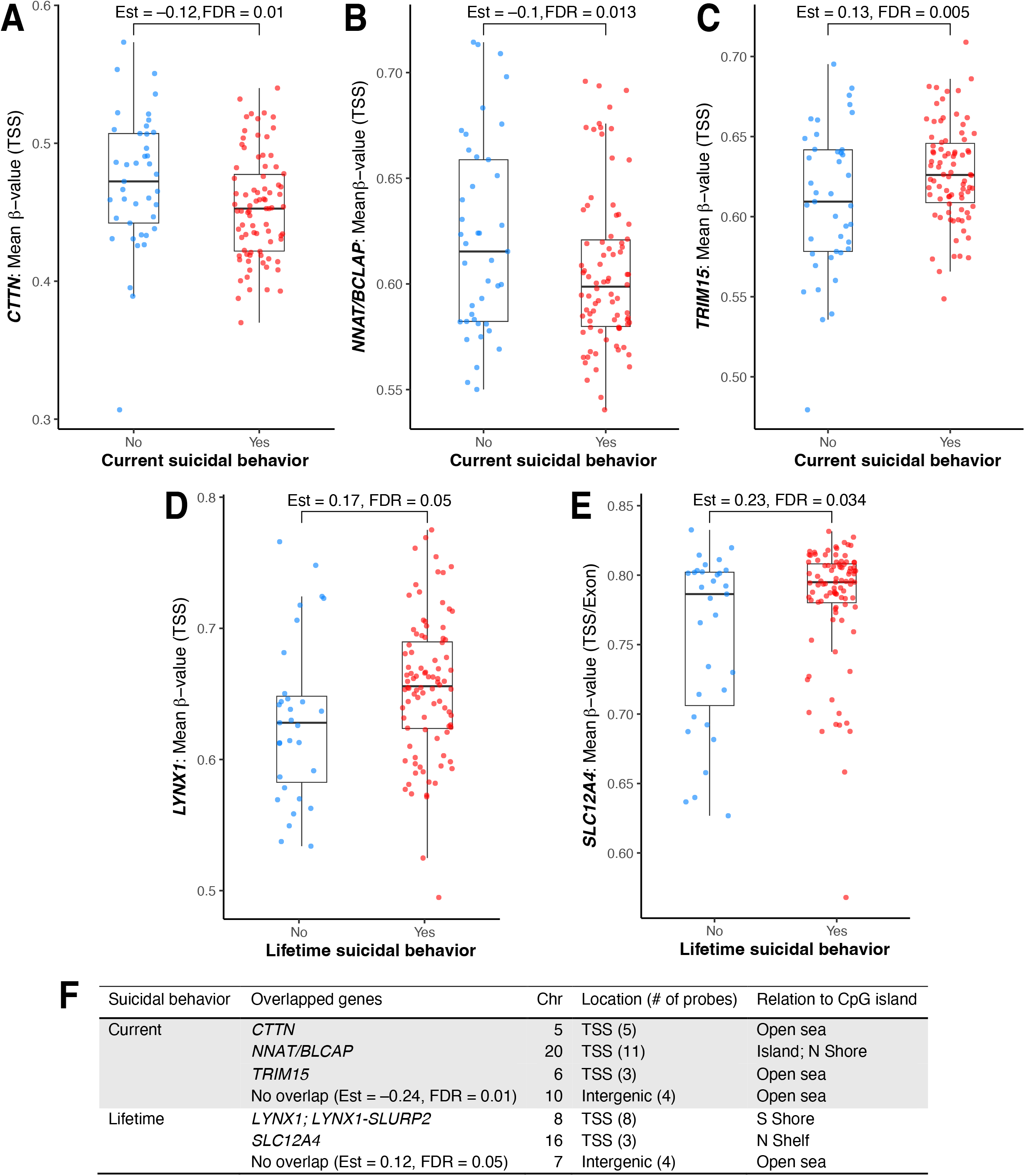
Differentially methylated genomic regions between individuals with and without current or lifetime suicidal behavior. Figures were generated from normalized β-values, statistical analyses were conducted using M-values. Averaged β-values among probes from regions related to the following genes: **A**: *CTTN*; **B**: *NNAT/BLCAP*; **C**: *TRIM15*; **D**: *LYNX1* **E**: *SLC12A4*. **F**: Genomic information of overlapped genes and DMR with no overlap to known genes. Relation to CpG island: TSS: 200-1,500 bp or 0-200 bp upstream of a TSS; CpG islands: CG dinucleotide-rich regions. N shore, S shore: 2,000 bp, directly up (Northern) and downstream (Southern) of CpG islands. N shelf, S shelf: 2,000 bp directly adjacent to shores; Opensea: methylation sites outside above-described regions.

**Figure 2.**
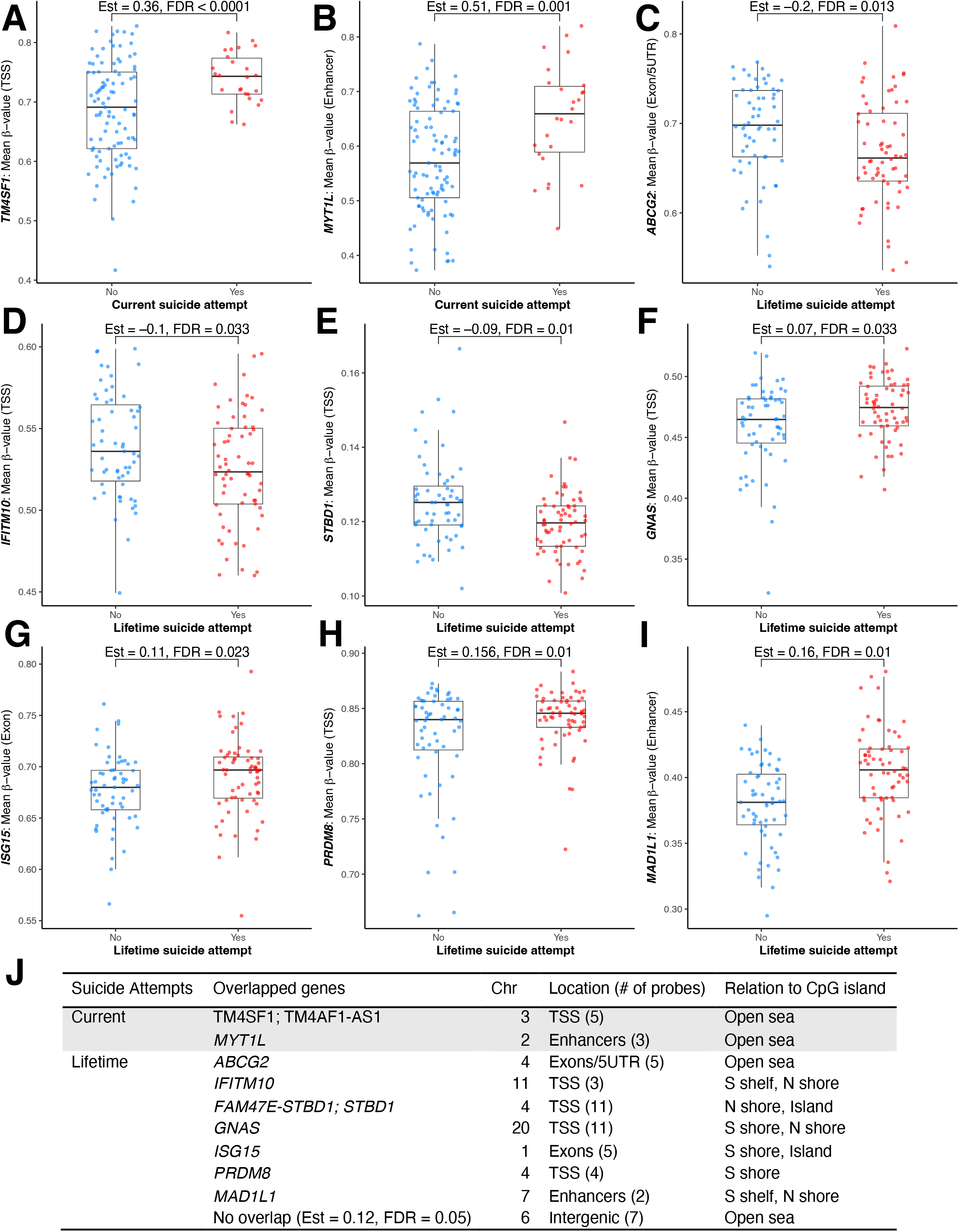
Differentially methylated genomic regions between individuals with and without current or lifetime suicide attempt. Figures were generated from normalized β-values, statistical analyses were conducted using M-values. Averaged β-values among the probes related to regions the following genes: **A**: *TM4SF1*; **B**: *MYT1L;* **C**: *ABCG2*; **D**: *IFITM10;* **E**: *STBD1;* **F**: *GNAS;* **G**: *ISG15;* **H**: *PRDM8;* **I**: *MAD1L1*. **J:** Genomic information of overlapped genes and DMRs with no overlap to known genes. Relation to CpG island as in Figure 1.

Next, we compared the average methylation levels of these genes, between groups with and without suicidal behavior or attempt, by including the probes located in the TSS, exon or enhancer regions. We found that methylation of *CTTN, NNAT/BLCAP* and *TRIM15* were significantly altered in subjects with current suicidal behavior and of *LYNX1* and *SLC12A4* in lifetime suicidal behavior (**Fig. 1**). For subjects that had actively attempted suicide, *TM4SF1* and *MYT1L* were significantly altered when the suicide attempt was recent (past 7 days), while *ABCG2*, *IFITM10*, *STBD1*, *GNAS*, *ISG15*, *PRDM8*, and *MAD1L1* were significantly altered in those with a lifetime history (**Fig. 2**).

### 2. Childhood trauma predicts lifetime suicide attempts

To determine if childhood trauma associated with suicidal behavior in adulthood in our cohort, we employed regression models using suicidal behavior and attempt, current and lifetime as the response variables and the ACE trauma-related questions as the independent variables, adjusting for sex, age and BMI. We found that the question ACE2, physical abuse in childhood, “*Did a parent or other adult in the household often push, grab, slap, or throw something at you? Ever hit you so hard that you had marks or were injured?”* was a significant predictor of suicide attempts during the subject’s lifetime (N: yes = 50, no = 61, *P* = 0.01). Subjects who experienced physical abuse in childhood had around 350% higher odds (95% CI: 40–1,320%) of a lifetime suicide attempt than those who did not.

Next, we performed a differential methylation analysis between subjects with and without physical abuse in childhood and identified 46,153 DMLs and seven DMRs overlapping seven unique genes (**Fig. 3A-F**). The *NNAT/BLCAP* gene pair, which overlapped with one of these DMRs, was also found to be directly associated with current suicidal behavior in the previous analysis. Additionally, three out of the four genes (*ADCY7, NNAT, FMOD, HOXA5*) have previously been linked to depression (**Supplementary Table 2**). Two of these genes (pairs) were hypomethylated (**Fig. 3A-B**) and four of them were hypermethylated in the group with childhood physical abuse (**Fig. 3C-F**). The detailed DMR descriptions are shown in **Fig. 3**. The identified genes related to suicidal behavior and childhood trauma together with brief function and involvement in mental health are presented in **Supplementary Table 2**.

**Figure 3.**
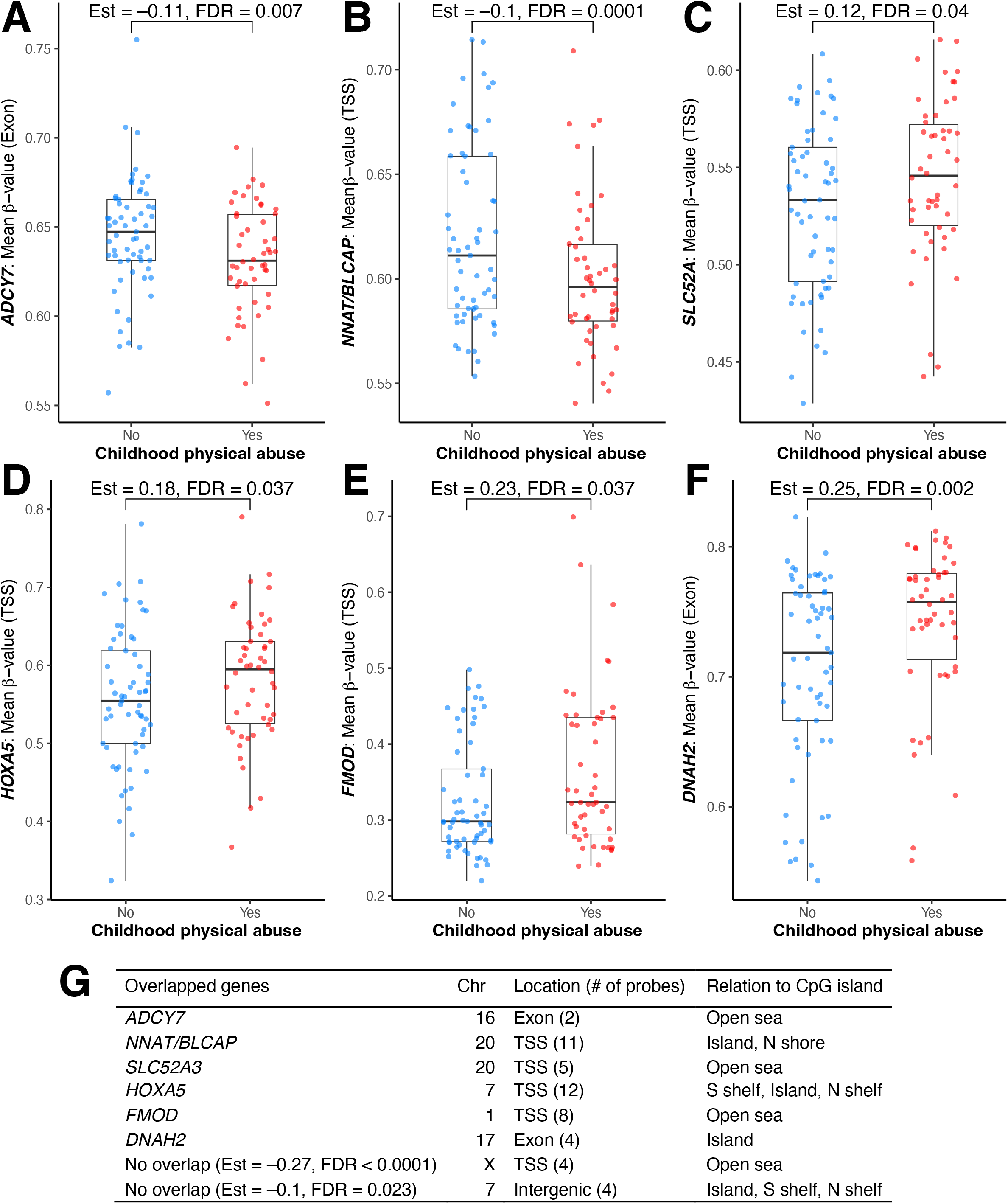
Differentially methylated genomic regions between individuals with and without childhood physical abuse trauma (ACE2). Figures were generated from normalized β-values, and statistical analyses were conducted using M-values. Averaged β-values from regions among the probes for each gene between groups with and without childhood physical abuse; **A**: *ADCY7*; **B**: *NNAT/BLCAP*; **C**: *SLC52A*; **D**: *HOXA5*; **E**: *FMOD*; **F**: *DNAH2*. **G.** Genomic information of overlapped genes and DMRs with no overlap to known genes between individuals with and without ACE2 trauma. N (yes = 50, no = 61). Relation to CpG island as in Figure 1.

### 3. Methylation predicts the HAMD suicide item dose-dependently

To further validate that the methylation patterns of the genes identified above were linked to suicidality, we conducted an ordinal regression analysis utilizing the HAMD item number 3 (severity of suicidality using discrete values from 0 to 4). We confirmed that the methylation levels of five of these genes significantly predicted HAMD-3 score using an ordinal regression model adjusted for sex, age and BMI (**Supplementary Table 3).**

### 4. Plasma immunometabolic biomarkers linked to future suicide attempts

We next conducted Bayesian logistic regression to determine whether a set of immunometabolic markers previously found to be linked to suicidal behavior (37) could predict suicide attempts over the year post enrollment. Serotonin had a strong protective effect, with lower levels predicting future suicide attempt (OR: 0.58, 95% CI: 0.39 to 1.13, posterior probability = 0.99) (**Supplementary Fig. 2A**). A two-fold nanomolar increase of serotonin was associated with a 31.4% decrease in the likelihood of a future suicide attempt. Thus, lower serotonin in plasma at baseline associated with suicide attempts over the year after initial enrollment (n=19). In contrast, there was no predictive effect of the tryptophan metabolites or cytokines measured (data not shown). We compared cell populations, using blood methylation data to deconvolute CD4 T-cells, CD8 T-cells, natural killer cells, B-cells, monocytes and neutrophils. Using Dirichlet regression for compositional data, we found that increased neutrophils in blood at baseline also was a significant predictor of suicide attempt over the next year (**Supplementary Fig. 2B**).

### 5. Prediction of suicide-attempts integrating methylation and immunometabolic factors

From the contrasts used to identify methylation patterns that distinguished individuals with suicidal behavior or attempts reported above, we identified 969 CpG sites that were shared among the four sets of DMLs (identifying current and lifetime suicidal behavior and attempts). Thus, we hypothesized that these individual CpG sites represent the most stable methylation markers associated with suicide risk. To narrow down these predictors, we first performed LASSO regression analysis, fitting all 969 CpG sites into the model to evaluate their predictive potential. The analysis identified 32 CpG sites with non-zero coefficients, suggesting that their contribution was significant. Of the 32 CpG sites, 11 were associated with known genes (annotation described in **Supplementary Table 4**). We confirmed that the direction of estimates for each CpG site was consistently positive or negative across the contrasts (estimate=Yes–No for each outcome; **Supplementary Table 5**), showing that the methylation levels were consistently higher or lower in the group with suicidal behavior or attempt.

To generate pilot prediction models, assessing the risk of future suicide attempts over a year, we utilized baseline methylation information (from both CpG sites and significant genes) and immunometabolic biomarkers linked with future suicide attempts. To future filter the 32 CpG sites, we performed PCA. The top two PCs clearly separated groups with and without future suicide attempt (**Supplementary Figure 3**), thus PC1 was used as a predictor. We first built a model using PC1, derived from the 32 CpG sites. We then built a second model using the top gene predictors: *STBD1*, *PRDM8,* and *TRIM15*, selected by ridge regression with the highest coefficients. In each model we included serotonin levels, neutrophil percentage, and confounders (sex, age, and BMI) to predict future suicide attempts. We evaluated performances of these models by assessing their discriminatory ability using ROC-AUC and PRC-AUC (**Fig. 4A-B**). The CpG-model had the best ROC-AUC (97%) and PRC-AUC (89%), followed by the gene-model (84% ROC-AUC and 54% PRC-AUC). **Fig. 4C** summarizes the models’ performance. We then performed Leave-one-out cross-validation, demonstrated that all alternative models had expected log predictive densities lower than the CpG-model (**Supplementary Table 6**). Posterior parameter estimates for this model are provided in **Supplementary Table 7**.

**Figure 4.**
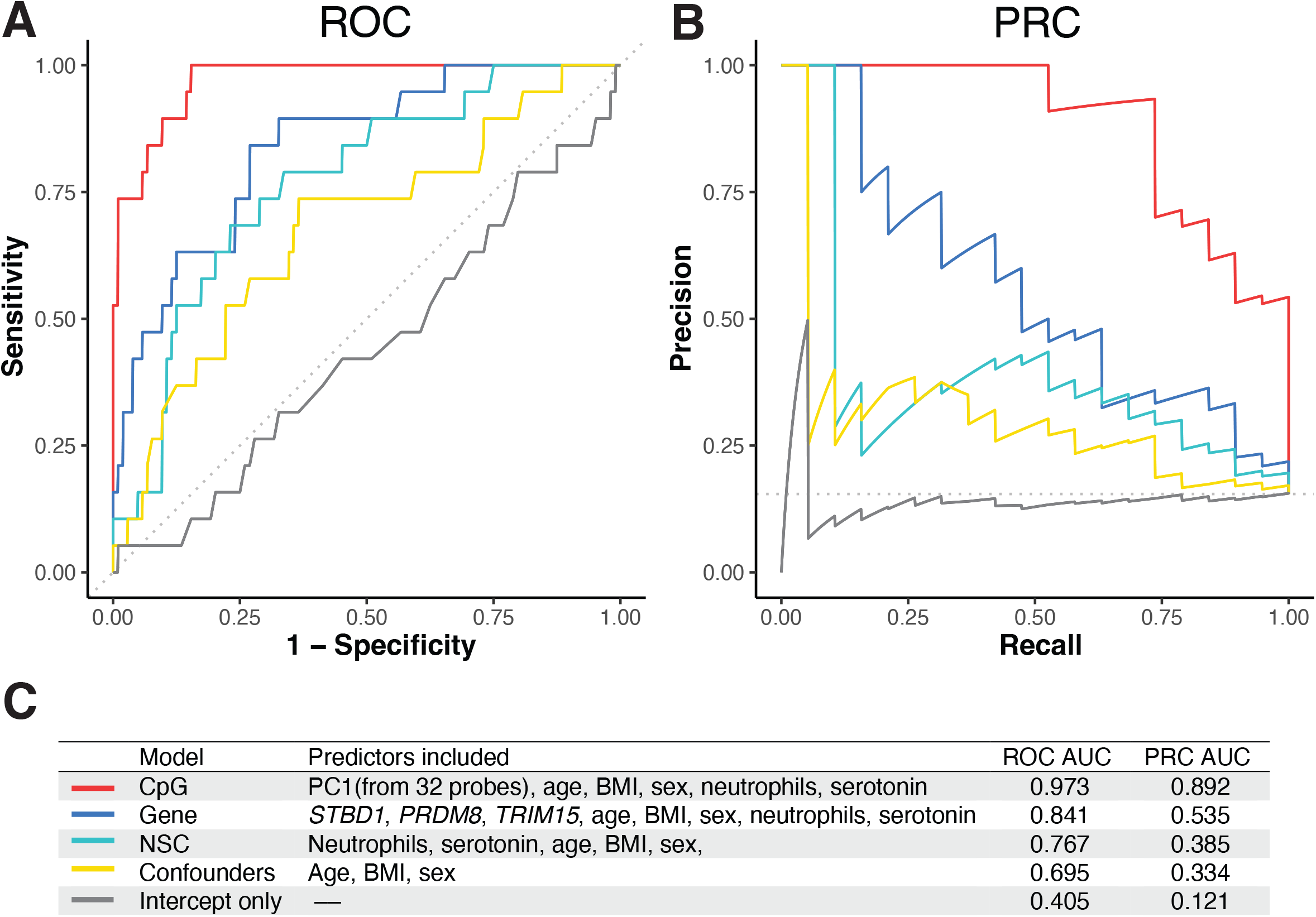
ROC-AUC and PRC-AUC from the models predicting future suicide attempts. **A**: Receiver Operating Characteristic AUC; **B:** Precision-Recall AUC. **C:** Prediction model descriptions with AUCs via Bayesian logistic regression with a horseshoe prior. Model adjusted for sex, age and BMI.

## Discussion

In this study, we enrolled 130 individuals with MDD, over half of whom presented with active suicidal behavior at intake. We found that the methylation pattern of 15 genes differed significantly based on current– or lifetime suicidal behavior, as determined at baseline. Moreover, childhood trauma predicted suicide attempts over a subjects’ lifetime and was linked to methylation changes in seven genes. We next used our data gathered at baseline to attempt to develop a preliminary prediction model that could indicate what patients, among the entire cohort of patients with moderate to severe depression, that were at increased risk for suicide attempts over the next 12 months. Lower levels of serotonin and increased neutrophil percentage were significant predictors of future suicide attempts in this population. Adding epigenetic marks from a gene model and a CpG-site model both increased the prediction accuracy. Our findings have clinical implications, as these biomarkers could potentially be used to identify MDD individuals who are at heightened risk for a suicide attempt in the intermediate timeframe (months to a year). Identifying this group allows for increased targeted preventative efforts and thereby potentially reduce risk of future suicidal behavior. This is the first report combining methylation patterns and immunometabolic blood biomarkers to predict future suicide attempts, highlighting novel possibilities to improve preventative patient care in clinical mental health settings.

Blood biomarkers may be useful in the assessments of suicide risk because they capture biological processes that reflect vulnerability to stress, psychiatric illness, and suicidal states in an objective and quantifiable way. Unlike self-reported questionaries or clinical interviews, blood biomarkers are not affected by stigma, insight, or unwillingness to disclose, and can be measured repeatedly over time, making them well-suited for individual longitudinal risk monitoring. Importantly, since no single biomarker is likely to be sufficiently predictive alone, combining immunometabolic markers and methylation signatures may better help identify individuals at elevated risk. Some biomarkers are likely to be more stable over time while others may fluctuate more acutely. For example, several studies (10, 12, 38) including this one, have found that methylation changes associated with childhood trauma are linked to future suicide risk –in our case *NNAT/BLCAP* was linked to both childhood trauma and suicidal behavior in later life– illustrating the usefulness of this type of biomarkers as they might be stable over many years.

Notably, several of the genes associated with suicidal behavior in our study have previously been linked to suicidality (**Supplementary Table 2**), strengthening the validity of the current data set. Among the genes we identified, in particular *MAD1L1* has repeatedly been associated with suicides and suicide attempts (39–42). *MAD1L1* encodes for MAD1, a mitotic spindle-assembly checkpoint protein, which was identified as a significant locus in a longitudinal EWAS study of depression (40). The authors identified a CpG site associated with *MAD1L1* that was significantly less methylated in the blood from individual with violent suicide attempts (n=31) than in those with non-violent attempts (n=57). Additionally, this CpG site was also hypermethylated in glial cells from depressed individuals in the validation cohort (40). In a recent study by Kim et.al., the authors used the data from two genetic consortia and multi-trait analysis of GWAS, identifying 114 polygenic risk loci for suicide attempt with one of them being localized with the intron region of *MAD1L1* (rs34021847) (42). Moreover, the methylation of *MAD1L1* was recently found to be altered in brain tissue from individuals with PTSD (43). In our study, we found two hypermethylated loci in an enhancer region of *MAD1L1* in individuals with a lifetime history of suicide attempts (**Fig. 2**). In the central nervous system, MAD1 is upregulated during active cortical development, and its deficiency leads to impairments in neuronal migration and neurite outgrowth (44). MAD1 interacts with KIFC3, a kinesin-like protein, regulating Golgi apparatus morphology and neuronal polarity to mediate proper neuronal migration and differentiation (44). Further studies to investigate the role of *MAD1L1* are highly warranted.

An additional four genes identified here have previously been reported linked to suicidal behavior. Kim et.al. reported lower expression of *TM4SF1* (a tetraspanin family cell-surface antigen) post-mortem prefrontal cortex tissue from suicide decedents diagnosed with bipolar disorder (45). In the same study, expression of *ABCG2*, encoding a plasma membrane efflux transporter, was higher in suicide decedents with schizophrenia (45). In our current study, we found that *TM4SF1* was hypermethylated in current suicide attempters, while *ABCG2* was hypomethylated in the group with lifetime suicide attempts. Additionally, using GWAS data, *Sokolowski et al.* showed that *GNAS,* encoding a G protein alpha subunit, was robustly associated with suicide attempts (46) and we reported differential methylation of *GNAS* in postmortem brain tissue from suicide decedents (1). In line with these findings, our current study shows *GNAS* hypermethylation in individuals with lifetime suicide history. Finally, *NINJ2*, encoding ninjurin 2, a homophilic transmembrane adhesion molecule, has been previously implicated in suicidality, with two SNPs within the *NINJ2* gene associated with suicide attempt in an Iranian population (47). Here, a CpG locus associated with *NINJ2* was strongly associated with future suicide attempts.

Given that physical trauma in childhood was a significant predictor of future suicide attempts, we examined whether there was overlap between genes with altered methylation linked to childhood trauma and genes linked to suicidal behavior in our cohort. Interestingly, one pair of genes, *NNAT/BLCAP,* was significantly altered in subjects with a history of childhood physical abuse and in individuals with suicidal behavior. Because the *NNAT* gene is located within an intron of the larger gene *BLCAP,* these genes are regulated together and usually denoted as a gene pair. *NNAT* encodes for neuronatin, an evolutionarily conserved protein expressed during brain development, especially in the fetal and neonatal brain. It is involved in calcium homeostasis and neuronal excitability. Low levels of neuronatin and genetic variants of *NNAT* have been linked with anorexia nervosa (48, 49). Also, a miRNA that regulates *NNAT* was recently found to be altered in patients with bipolar disorder and linked to depressive-like behavior in mice (50).

As the final part of this study, we performed an exploratory prediction analysis using ROC to determine whether our immunometabolic and epigenetic datasets were associated with suicide attempts over the year after enrollment. Interestingly, we found that low blood serotonin and increased neutrophil percentage at baseline predicted suicide attempts in our study. Both these factors have previously been linked to current suicidal behavior and attempts in cross-sectional studies (51, 52). We therefore propose that when used together with clinical data, the epigenetic and immuno-metabolic blood biomarkers identified here, could improve risk stratification and support earlier intervention, advancing suicide prevention towards a more personalized and biologically informed approach over an intermediate time frame (weeks to a year after evaluation). However, we did not detect predictive effects for tryptophan, the panel of kynurenine metabolites, or cytokines measured here. Such rapidly fluctuating biomarkers may instead reflect changes in suicide risk in the more acute time frame of days (53), warranting further studies moving forward.

In contrast to previous studies on suicidal behavior, which often include participants with various psychiatric disorders, our study was designed to focus on a diagnostically homogenous population of individuals diagnosed with moderate to severe MDD, stratified by the presence or absence of suicidal behavior. Subjects were confirmed to be free of immunomodulatory medications, potential confounding inflammatory conditions or ongoing infections. Although this strategy enhanced the likelihood of detecting changes specifically related to suicidal behavior, the extent to which the findings are generalizable warrants consideration. Importantly, many of the identified genes have previously been linked to psychiatric disease, indicating that the observed effects extend beyond this specific cohort. Similarly, lower levels of serotonin in peripheral blood, and increased inflammatory status in the form of higher neutrophil to lymphocyte ratio have been consistently observed in suicidal cohorts (54–57). Altogether, several of the genes and markers identified here validate findings from other cohorts, highlighting the robustness of our results and their potential relevance beyond our sample.

## Conclusions

We identified significantly altered DNA-methylation in MDD individuals with suicidal behavior and in those who had experienced childhood trauma. We also developed models to predict future suicide attempts over the post-enrollment year that included the epigenetic markers, increased neutrophil percentage in blood and lower plasma serotonin levels at baseline. Importantly, our timeframe can be useful for future preventative efforts in clinical settings, as it could indicate which patients need a heightened level of follow-up and care after discharge. The neurobiological mechanisms linked to these immunometabolic and epigenetic changes also warrant further study to improve our understanding of suicidal behavior.

## Data Availability

All data produced in the present study are available upon reasonable request to the authors

## Acknowledgements

The study was funded by the National Institutes of Mental Health (NIMH R01 MH118211) awarded to the multi-PI team of Drs Brundin, Mann and Achtyes. We wish to thank all the participants for their participation in this study.

