## Supplemental materials for "Epigenetic and Immunometabolic Signatures of Suicidal Behavior in Major Depressive Disorder"

**Supplementary materials**

**Supplementary Figure 1**

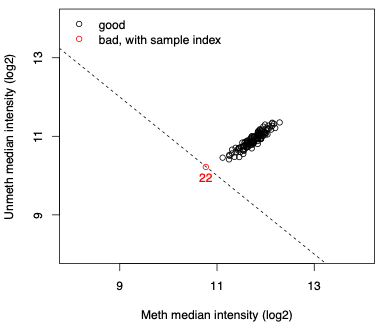

**Supplementary Figure 1. Quality control plot of the methylation data on 124 samples.** One out of 124 samples was identified as an outlier, thus removed for future analysis. The plot was generated based on joint distributions of log₂-transformed median methylated and unmethylated signal intensities, visualized using plotQC function within the minfi package.

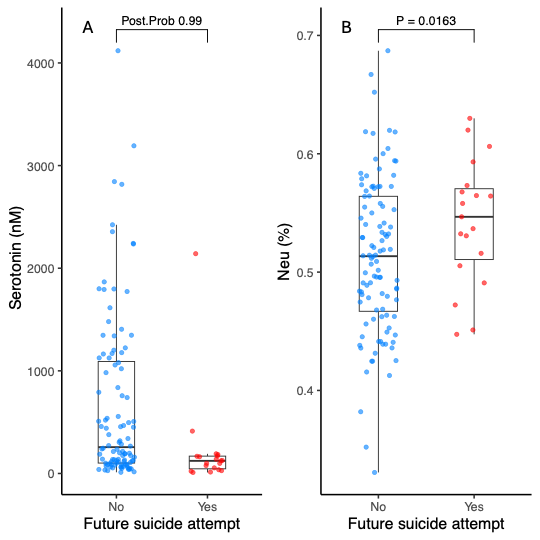

**Supplementary Figure 2.** **Serotonin levels and percentage of neutrophils differ between individuals who attempted suicide over the year following discharge and those who did not.** **A:** Serotonin level difference. Bayesian logistic regression with a horseshoe prior adjusting for sex, age, and BMI. OR: 0.58, 95% CI: 0.39–1.13. **B:** Neutrophil population difference. Dirichlet regression, adjusting for sex, age and BMI, was used to compare the differences between groups with and without future suicide attempt for neutrophil percentage. Estimate = 0.42.

| Category | Current suicidal behavior | | Suicide attempt within a year post-enrollment | |
| --- | --- | --- | --- | --- |
|  | Yes  (N = 82) | No  (N = 41) | Yes  (N = 19) | No  (N = 104) |
| SSRI/SNRI (antidepressant) | 74 (90%) | 33 (81%) | 18 (95%) | 89 (86%) |
| Antipsychotic | 34 (42%) | 6 (15%) | 13 (68%) | 27 (26%) |
| Antihistamines | 28 (34%) | 21 (51%) | 7 (37%) | 42 (40%) |
| Anti-inflammatories | 26 (32%) | 12 (29%) | 7 (37%) | 31 (30%) |
| Antiepileptics | 22 (27%) | 3 (7%) | 7 (37%) | 18 (17%) |
| Atypical-antidepressants | 15 (18%) | 6 (15%) | 16 (84%) | 16 (15%) |
| Proton pump inhibitors | 10 (12%) | 3 (7%) | 9 (47%) | 9 (9%) |
| Lipid lowering | 8 (10%) | 8 (20%) | 13 (68%) | 13 (13%) |
| Anxiolytic (non-benzodiazepine) | 7 (9%) | 7 (17%) | 12 (63%) | 12 (2%) |

**Supplementary Table 1.** Main medication categories by the participants. Total N = 123. Current suicidal behavior was identified by “yes” to C-SSRS 21 assessed over the prior 7 days at enrollment. Future suicide attempt was identified by “yes” to either C-SSRS13 or C-SSRS16 over the year after discharge from the hospital.

**Supplementary Table 2.** Reported links to mental health disorders of the differentially methylated genes that were identified in this study. Grouped by the clinical characteristics/analysis of the group in which they were identified.

|  | **Gene(s)** | **Function related to mental health** |
| --- | --- | --- |
| Current suicidal behavior and childhood trauma physical abuse | *NNAT/BLCAP* | Neuronatin. Proteolipid that may be involved in the ion channels regulation during brain development (1) Bladder Cancer Associated Protein. Protein that reduces cell growth by stimulating apoptosis. NNAT is linked to depressive-like behavior in mice (2). |
|  | *CTTN* | Encodes cortacin. Cortacin expression was decreased the postmortem brain tissue in patients with schizophrenia versus control (3). |
| Lifetime suicidal behavior | *LYNX1* | Ly6/Neurotoxin 1. Membrane-bound protein that binds to nicotinic acetylcholine receptors (nAChRs) in the mammalian brain, acting as a “brake” on synaptic plasticity (1). Differentially methylated in depressed pregnant women (4, 5) |
| Current suicide attempt | *MYT1L* | Myelin Transcription Factor 1 Like. a neural-specific transcription factor essential for brain development and the maintenance of neuronal identity (1). Risk gene for MDD (6). Copy number variation of this gene has been linked to schizophrenia (7) |
|  | *TM4SF1* | A tetraspanin family cell-surface antigen. Differentially expressed in the suicide completers from the Bipolar patients (8). |
| Lifetime suicide attempt | *IFITM10* | Interferon Induced Transmembrane Protein 10. A IFITM10 SNP been linked to the age of onset of schizophrenia (9). |
|  | *ABCG2* | Encodes a plasma membrane efflux transporter, belongs to ABC transporter. Differentially expressed in the suicide completers from the schizophrenia patients (8). |
|  | *GNAS* | Guanine Nucleotide Binding Protein (G Protein) Alpha. Differentially methylated in depressed pregnant women compared to controls [4]. In a GWAS study, *GNAS* was robustly associated with suicide attempt using different methods (10). It has shown differentially methylated in the postmortem brain tissue from the suicide completers (11). |
|  | *ISG15* | Interferon-Stimulated Gene 15. Functions as a ubiquitin-like modifier (ISGylation) to inhibit viral replication (12). Upregulating this gene causing causes depressive-like behavior in mice (13). |
|  | *PRDM8* | PR/SET Domain 8. Encodes a protein belongs to the family of histone methyltransferases, acts as negative regulator of transcription (1). Hypomethylated in the borderline personality disorder (14). |
|  | *MAD1L1* | Mitotic Arrest Deficient 1 Like 1. Component of the spindle-assembly checkpoint (1). Associated with suicide (15, 16) depression (15, 17) and schizophrenia (18). |
| Childhood trauma | *ADCY7* | Adenylate Cyclase 7. Encodes a membrane-bound adenylate cyclase that is inhibitable by calcium [1]. Implicated in MDD (19, 20) and alcohol use disorder (19). |
|  | *HOXA5* | Homeobox A5. Encodes transcription factors, part of A cluster on chromosome 7 (1). Differentially methylated in the depressed pregnant women compared to controls (5). |
|  | *FMOD* | Encodes fibromodulin, that belongs to the family of small interstitial proteoglycans, affects the rate of fibrils formation (1). Alleviates depression-like behavior in mouse models (21). |
| 32 CpG sites | *SELENBP1* | Selenium-binding protein, involved in neurological disorders including Parkinson’s, schizophrenia. Knockout of this gene in mouse induced depressive like behavior (22). |
|  | *TKT* | CutA Divalent Cation Tolerance Like, Pseudogene. Thiamine-dependent enzyme that catalyzes the metabolism of sugars, linking the pentose phosphate pathway to glycolysis (1). Loss of TKT activity, leads to Wernicke encephalopathy in alcoholics and abnormal eye movements and ataxia in children with thiamin deficiency (23). Thiamine deficiency is also linked to depression in older adults (24). |
|  | *NINJ2* | Nerve Injury-Induced Protein 2. Mice with oligodendrocyte-specific deletion of NINJ2 exhibit depressive-like behavior (25). SNPs within the *NINJ2* gene were associated with suicide attempt in an Iranian population (26). |
|  | *SLITRK5* | SLIT and NTRK Like Family Member 5. Expressed predominantly in neural tissues with demonstrated neurite-modulating activity. Involved in Obsessive-Compulsive Disorder, Attention-Deficit/Hyperactivity Disorder and Autism Spectrum Disorder (27). |
|  | *NFATC3* | Nuclear Factor of Activated T Cells 3. Regulation of gene expression in T cells and immature thymocytes, linked to schizophrenia Click or tap here to enter text.. Altered calcineurin / NFAT activation is linked to neurodegenerative diseases characterized by synaptic dysfunction, glial activation, and neuronal death (28). NFATC3 is significantly increased in schizophrenia and bipolar disorder (29) |

**Supplementary Table 3.** Genes significantly associated with HAMD suicide rating scale (item 3) by ordinal regression, adjusting for age, sex and BMI.

| Gene | estimate | P-value |
| --- | --- | --- |
| *TM4SF1* | 0.83 | 0.018 |
| *MYT1L* | 0.71 | 0.019 |
| *STBD1* | -4.14 | 0.003 |
| *PRDM8* | 1.05 | 0.037 |
| *NNAT/BLCAP* | -2.14 | 0.007 |

**Supplementary Table 4.** CpG sites associated with known genes selected from the LASSO model.

| Probe ID | Chr | Estimate  (Yes – No) | UCSC RefGene | UCSC RefGene Group | UCSC CpG Island |
| --- | --- | --- | --- | --- | --- |
| cg24355850_BC11 | 1 | Positive | *LINC01144* | TSS200 | Island |
| cg11194923_BC21 | 1 | Negative | *SELENBP1* | exon | N/A^1^ |
| cg24969254_BC21 | 3 | Negative | *TKT* | 3UTR; exon | N/A |
| cg09295209_BC21 | 3 | Positive | *LOC102724604* | TSS200 | N/A |
| cg11486912_BC21 | 9 | Positive | *CUTALP* | TSS1500 | S Shelf |
| cg20379954_TC21 | 12 | Negative | *NINJ2* | TSS200 | S Shore |
| cg07710481_BC21 | 13 | Negative | *SLITRK5* | TSS200 | S Shore; N Shore |
| cg08392069_TC21 | 16 | Positive | *SNHG19; TRAF7; SNORD60* | exon; TSS1500; TSS200 | N Shore |
| cg07981599_TC11 | 16 | Positive | *NFATC3* | TSS1500 | Island |
| cg27665181_TC11 | 16 | Negative | *MTSS2* | TSS200 | Island |
| cg26909237_BC21 | 18 | Positive | *FBXO15; TIMM21* | 5UTR; exon1; TSS1500 | Island |

1. Information not available.

**Supplementary Table 5.** DML results of the 32 probes in the four methylation analyses. Abbreviations: CSB, Current suicidal behavior; LSB, Lifetime suicidal behavior; CSA, Current suicide attempt; LSA, Lifetime suicide attempt.

| Probe_ID | Group  comparison | Estimate  (Yes – No) | P |
| --- | --- | --- | --- |
| cg24355850_BC11 | CSB | 0.28516332 | 0.0002459610 |
|  | LSB | 0.29297755 | 0.0004411927 |
|  | CSA | 0.21139148 | 0.0179966295 |
|  | LSA | 0.16605211 | 2.563516e-02 |
| cg11194923_BC21 | CSB | -0.07901525 | 0.0285504316 |
|  | LSB | -0.084842101 | 0.027910175 |
|  | CSA | -0.093025717 | 0.022424273 |
|  | LSA | -0.073022265 | 0.031362545 |
| cg01615585_BC21 | CSB | 0.075228471 | 0.009823775 |
|  | LSB | 0.073652051 | 0.018367409 |
|  | CSA | 0.094323864 | 0.004049675 |
|  | LSA | 0.086041937 | 0.001553667 |
| cg03132634_BC21 | CSB | -0.082162547 | 0.045440659 |
|  | LSB | -0.091205472 | 0.037723498 |
|  | CSA | -0.111027196 | 0.01634808 |
|  | LSA | -0.099820937 | 0.009285969 |
| cg03062942_BC21 | CSB | 0.107272068 | 0.02002596 |
|  | LSB | 0.116108853 | 0.018543895 |
|  | CSA | 0.138980662 | 0.007443954 |
|  | LSA | 0.120982032 | 0.005021256 |
| cg05127937_BC21 | CSB | -0.064503613 | 0.009839644 |
|  | LSB | -0.053341114 | 0.047194817 |
|  | CSA | -0.075644961 | 0.007324368 |
|  | LSA | -0.052634617 | 0.025520848 |
| cg11753709_BC21 | CSB | -0.076437236 | 0.009281712 |
|  | LSB | -0.075150808 | 0.017026551 |
|  | CSA | -0.101169099 | 0.002186661 |
|  | LSA | -0.078607037 | 0.004303449 |
| cg24969254_BC21 | CSB | -0.113198288 | 7.71E-04 |
|  | LSB | -0.115630381 | 0.00135028 |
|  | CSA | -0.13341518 | 4.40E-04 |
|  | LSA | -0.083639338 | 0.008809241 |
| cg05965863_TC21 | CSB | -0.098720535 | 0.02957948 |
|  | LSB | -0.11209766 | 0.020722535 |
|  | CSA | -0.124800639 | 0.014684804 |
|  | LSA | -0.104043225 | 0.014432718 |
| cg05606728_BC21 | CSB | -0.164430889 | 9.30E-05 |
|  | LSB | -0.104432688 | 0.023079242 |
|  | CSA | -0.12979945 | 0.007273799 |
|  | LSA | -0.094798604 | 0.018830633 |
| cg22504438_BC21 | CSB | -0.091282702 | 0.004855532 |
|  | LSB | -0.092504605 | 0.007720678 |
|  | CSA | -0.079670105 | 0.030756573 |
|  | LSA | -0.073624975 | 0.016116845 |
| cg09295209_BC21 | CSB | 0.112125509 | 0.019321681 |
|  | LSB | 0.103727273 | 0.043646374 |
|  | CSA | 0.150714574 | 0.005171948 |
|  | LSA | 0.123123158 | 0.006048042 |
| cg07162914_BC21 | CSB | 0.11780311 | 0.032786265 |
|  | LSB | 0.125367093 | 0.033673375 |
|  | CSA | 0.17088344 | 0.005820676 |
|  | LSA | 0.136512689 | 0.00815011 |
| cg10042397_TC21 | CSB | 0.1331578 | 0.01160287 |
|  | LSB | 0.136320994 | 0.01580891 |
|  | CSA | 0.131437438 | 0.027993734 |
|  | LSA | 0.131207984 | 0.008049662 |
| cg07332511_TC21 | CSB | 0.074495169 | 0.02730536 |
|  | LSB | 0.100808327 | 0.004945187 |
|  | CSA | 0.077182958 | 0.043373961 |
|  | LSA | 0.111276135 | 3.62E-04 |
| cg11486912_BC21 | CSB | 0.101080088 | 0.003260131 |
|  | LSB | 0.115597692 | 0.001607055 |
|  | CSA | 0.082086552 | 0.036151076 |
|  | LSA | 0.082449354 | 0.011030198 |
| cg10266121_TC21 | CSB | 0.076156119 | 0.020742374 |
|  | LSB | 0.114824209 | 9.72E-04 |
|  | CSA | 0.074366995 | 0.046317987 |
|  | LSA | 0.099351501 | 0.001172463 |
| cg24616385_TC21 | CSB | 0.131537237 | 0.002595514 |
|  | LSB | 0.145133379 | 0.001862178 |
|  | CSA | 0.129700109 | 0.008905465 |
|  | LSA | 0.149056349 | 2.47E-04 |
| cg20379954_TC21 | CSB | -0.118946277 | 0.049780102 |
|  | LSB | -0.149601489 | 0.020638666 |
|  | CSA | -0.206388726 | 0.002312256 |
|  | LSA | -0.139242476 | 0.014088958 |
| cg18147331_TC21 | CSB | -0.206877019 | 0.001965269 |
|  | LSB | -0.229117711 | 0.001325643 |
|  | CSA | -0.178705508 | 0.018985769 |
|  | LSA | -0.241740199 | 9.86E-05 |
| cg13734792_BC21 | CSB | 0.070104958 | 0.015215514 |
|  | LSB | 0.061469853 | 0.047509803 |
|  | CSA | 0.074996942 | 0.021756774 |
|  | LSA | 0.054442524 | 0.045757191 |
| cg07710481_BC21 | CSB | -0.135039816 | 0.022106051 |
|  | LSB | -0.159010602 | 0.011543872 |
|  | CSA | -0.154337137 | 0.020611375 |
|  | LSA | -0.140955151 | 0.010817029 |
| cg02633600_TC21 | CSB | 0.122624994 | 0.001951923 |
|  | LSB | 0.109231923 | 0.010361233 |
|  | CSA | 0.09522839 | 0.035252441 |
|  | LSA | 0.097686148 | 0.009047929 |
| cg08392069_TC21 | CSB | 0.100460777 | 0.033285911 |
|  | LSB | 0.111229438 | 0.027438425 |
|  | CSA | 0.118039533 | 0.026768405 |
|  | LSA | 0.125965222 | 0.004209878 |
| cg07981599_TC11 | CSB | 0.295628005 | 6.66E-04 |
|  | LSB | 0.23899343 | 0.010925952 |
|  | CSA | 0.270066198 | 0.006388112 |
|  | LSA | 0.171853946 | 0.038168751 |
| cg27665181_TC11 | CSB | -0.097237606 | 0.017886474 |
|  | LSB | -0.110853888 | 0.011459892 |
|  | CSA | -0.105358001 | 0.023323833 |
|  | LSA | -0.076300825 | 0.048848666 |
| cg26909237_BC21 | CSB | 0.142909724 | 8.78E-04 |
|  | LSB | 0.178938428 | 8.46E-05 |
|  | CSA | 0.102460548 | 0.037432241 |
|  | LSA | 0.089168117 | 0.029278144 |
| cg16385423_TC21 | CSB | 0.092925015 | 0.012837483 |
|  | LSB | 0.124336775 | 0.001706484 |
|  | CSA | 0.102790952 | 0.014926489 |
|  | LSA | 0.104384566 | 0.002776531 |
| cg01408926_TC21 | CSB | 0.061269142 | 0.01203111 |
|  | LSB | 0.069313671 | 0.007797464 |
|  | CSA | 0.067761841 | 0.014043112 |
|  | LSA | 0.056013928 | 0.014642634 |
| cg01602479_TC21 | CSB | -0.123288534 | 0.005812172 |
|  | LSB | -0.127239257 | 0.00784611 |
|  | CSA | -0.140996035 | 0.005233778 |
|  | LSA | -0.092032068 | 0.029435993 |
| cg12647020_TC21 | CSB | -0.081797743 | 0.023170522 |
|  | LSB | -0.080844856 | 0.036174258 |
|  | CSA | -0.081020694 | 0.04714578 |
|  | LSA | -0.07276985 | 0.031800336 |
| cg22332233_BC21 | CSB | -0.091595361 | 0.037128415 |
|  | LSB | -0.119068778 | 0.010929547 |
|  | CSA | -0.09836439 | 0.047830201 |
|  | LSA | -0.081840279 | 0.047718113 |

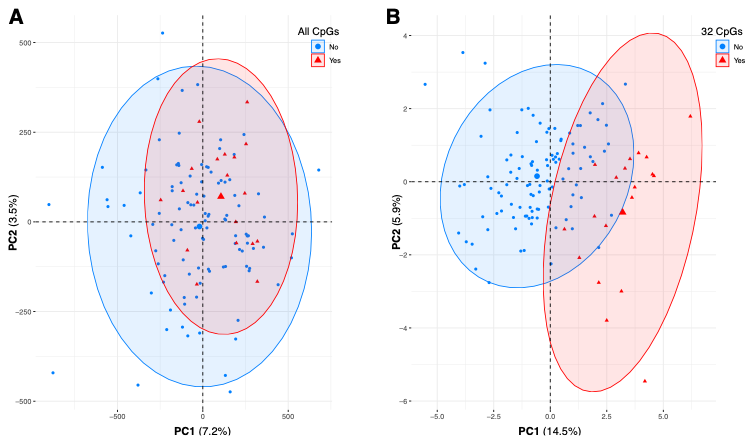

**Supplementary Figure3 Principal Component Analysis.** PCA was performed on the M-values of each of the total 900,272 CpGs probes measusred (A) or the selected 32 CpGs probes by LASSO regression (B) at baseline. Each point represents an individual; individuals with reported suicide attempts within a year post enrollment are shown in red (N=19) while participants without suicide attempts are in blue (N=104). In A, the first two principal components explain 7.2% (PC1) and 3.5% (PC2) of the total variance and samples did not cluster by group. In B, the first two principal components explain 14.5% (PC1) and 5.9% (PC2) of the total variance; separation along PC1 indicates that the methylation status of the selected 32 CpGs at baseline can be used to distinguish individuals who will attempt suicide in the future (group = Yes).

**Supplementary Table 6.** Model comparisons by LOO.

| Ranking | Model | ELPD diff | SE diff |
| --- | --- | --- | --- |
| 1 | PC1, probes, serotonin, neutrophils, confounders | 0.0 | 0.0 |
| 2 | 3 top genes, serotonin, neutrophils, confounders | -23.3 | 5.7 |
| 3 | Serotonin, neutrophils, confounders | -25.8 | 5.7 |
| 4 | Only confounders | -30.0 | 6.4 |
| 5 | Intercept | -30.2 | 6.4 |

Notes: ELPD diff: Expected Log Predictive Density difference between each model and the best model; SE diff: standard error of ELPD difference between each model and the best model.

**Supplementary Table 7.** Output from Bayesian’s logistic regression model, using a horseshoe prior. Outcome: future suicide attempt.

| Model | Predictors included | Estimate (95% CI) |
| --- | --- | --- |
| CpG model | PC1(from 32 probes), serotonin, neutrophils, confounders (age, BMI, sex) | PC1: 1.53 (1, 2.15);  Age: -0.03(-0.13, 0.04);  SexM: -0.27 (-1.81, 0.68);  BMI: 0.02 (-0.06, 0.12);  Neu: 0.05 (-1.8, 1.99);  Serotonin: -0.3 (-1.03, 0.18) |
| Gene model | STBD1, PRDM8, TRIM15, serotonin, neutrophils, confounders (age, BMI, sex) | TRIM15: 0.79 (-0.33, 2.82);  STBD1: -0.87 (-3.74, 0.45);  PRDM8: 1.05 (-0.06, 2.76);  Age: -0.04 (-0.1, 0.01);  SexM: -0.78 (-2.35, 0.2);  BMI: 0.01 (-0.05, 0.07);  Neu: 0.45 (-1.27, 3.77);  Serotonin: -0.64 (-1.18, -0.13) |
| Serotonin/  Neutrophils | Serotonin, neutrophils, confounders (age, BMI, sex) | Age: -0.04 (-0.09, 0.01);  SexM: -0.48 (-1.93, 0.23);  BMI: 0.01 (-0.04, 0.07);  Neu: 0.22 (-0.98, 2.43);  Serotonin: -0.54 (-1.04, -0.01) |

**References (supplemental):**

1.	Science WIo, GeneCards the human gene database.

8.	Kim S, Choi KH, Baykiz AF, HK. G (2007): Suicide candidate genes associated with bipolar disorder and schizophrenia: an exploratory gene expression profiling analysis of post-mortem prefrontal cortex. *BMC Genomics*. 12.
